# At-home validation of remote breathing monitoring: A proof-of-concept for long-term care of respiratory patients using a non-contact, radar-based biomotion sensor

**DOI:** 10.1101/2024.03.17.24304031

**Authors:** Tobit Fischer, Torsten Eggert, Alina Wildenauer, Sarah Dietz-Terjung, Rainer Voisard, Christoph Schöbel

**Author notes:** Correspondence: Christoph Schöbel Universitätsmedizin Essen – Ruhrlandklinik Tüschener Weg 40 45239 Essen, Germany.

## Abstract

**Purpose:** Long-term monitoring of respiratory rate (RR) is promising for the management of chronic conditions. Research interest is particularly high in chronic respiratory diseases (CRDs), especially for predicting acute exacerbations of COPD (AECOPD). The aim of the present study was to evaluate the long-term validity of a recent non-contact biomotion sensor in the home environment of CRD patients with domiciliary ventilator support, focusing on patient acceptance and usability of this device, as well as RR fluctuations related to AECOPD.

**Patients and methods:** In this prospective proof-of-concept study, 19 patients requiring non-invasive ventilation (NIV) and seven patients requiring invasive mechanical ventilation (IMV) were provided with the non-contact device for six and one month, respectively. Main indication for NIV therapy was COPD. Real-world validation of the device was performed by comparing nocturnal RR values between the non-contact system and both types of ventilators. The acceptance and operability of the biomotion sensor were evaluated using a questionnaire. COPD exacerbations that occurred during the study period were assessed for possible RR fluctuations preceding these events.

**Results:** Mean absolute error (MAE) of median RR between the NIV device and the non-contact system, based on 2326 nights, was 0.78 (SD: 1.96) breaths per minute (brpm). MAE between the IMV device and the non-contact system was 0.12 brpm (SD: 0.52) for 215 nights. The non-contact device was accepted by the patients and proved to be easy to use. In some of the overall 13 cases of AECOPD, RR time courses showed variations of increased nocturnal respiratory activity a few days before the occurrence of such events.

**Conclusion:** The present non-contact system is suitable and well accepted for valid long-term monitoring of nocturnal RR in the patient’s home environment. This finding may serve as a starting point for larger studies, e.g., to develop robust AECOPD prediction rules.

**KEY MASSAGES:** 

**What is already known on this topic:** Technological advances offer new possibilities for remote respiratory rate (RR) monitoring for various use cases. Although novel devices are regularly validated in an experimental environment, the often-recommended verification of this validity in long-term use, including an assessment of patient adherence, is lacking.

**What this study adds:** One of the main challenges is the absence of a feasible reference standard for long-term RR measurement. The present work demonstrates a new approach to validate a non-contact device in the home environment by comparing RR measurements with built-in software data in a cohort of ventilator-dependent patients.

**How this study might affect research, practice or policy:** Patient compliance, difficulties, as well as their needs and requirements for such long-term monitoring were recorded in order to improve further patient-centered studies. Following this premise, we aim to encourage the development of suitable validation standards, as long-term RR monitoring could finally become viable.

## INTRODUCTION

Technological innovations enable continuous monitoring and digital transmission of vital signs and disease-specific parameters to healthcare providers. The increasing acceptance of such telemedical services, accelerated by the COVID-19 pandemic, creates promising opportunities for personalized smart healthcare in chronic diseases.(1) In addition to the adequate validation of new technologies, active involvement from both patients and practitioners is crucial to gather practical insights, thereby achieving effective and well-accepted integration into the healthcare system. While respiratory rate (RR) is an integral part of clinical care and assessment, e.g., for pneumonia or sepsis,(2,3) ongoing research explores its utility beyond hospital settings for chronic conditions. For example, investigations include RR’s predictive role in chronic heart failure,(4) cystic fibrosis,(5) and chronic obstructive pulmonary disease (COPD).(6–9)

COPD is the most prevalent among chronic respiratory diseases (CRD), which are acknowledged as one of the top leading causes of death worldwide.(10,11) Over the past decade, considerable effort has been made in COPD research to improve disease management, with the focus on early detection and prevention of acute exacerbations of COPD (AECOPD), as they are known to have a negative impact on disease progression and the patient’s health-related quality of life.(12,13) Various studies suggest that there are fluctuations in nocturnal RR(6–9) occurring during a brief prodrome, which is known to precede COPD exacerbations.(14,15) Thus, early recognition of significant RR changes would allow for timely intervention, potentially reducing the need for hospitalization or leading to faster recovery from a worsening of symptoms.(16) This could be, for example, accomplished by the implementation of continuous RR monitoring in the home environment.(6–9)

Up to now, various attempts have been made to monitor RR at home, for example, via undergarment waistband-adhered physiologic monitors,(17) via monitors embedded in the domiciliary oxygen supply system,(9) by analyzing RR estimates from non-invasive ventilator software,(7,8) by minimal-contact sensors placed under the mattress,(18) or by using a non-contact biomotion sensor to measure thoracic excursions.(6,19,20) Home-based sleep sensors offer distinct advantages. Confounding factors like mental strain, heat, cold, or physical effort,(21) are expected to be minimized. Additionally, contactless approaches with non-impairing designs, coupled with automated data collection and transmission, may enhance compliance and enable extended data acquisition in familiar environments, facilitating trend analyses.

Novel devices designed to monitor vital parameters typically undergo initial validation in controlled laboratory settings for a limited usage time. Despite the intrinsic strength of these devices for extended monitoring in the home environment, re-evaluation over prolonged periods is rarely carried out. The main reason may be that the established reference methods for measuring RR, such as manually counting breaths or using a thoracic respiratory effort belt (TREB), are not well suited for long-term home monitoring. Along with the frequent lack of analysis of patient compliance, these factors may contribute to the fact that none of such devices have yet become widely accepted.

The present proof-of-concept evaluates the Sleepiz One+ non-contact biomotion sensor (Sleepiz AG, Zurich, Switzerland; hereafter referred to as contactless sleep monitor or CSM) as an outpatient approach to monitor nocturnal RR. Its RR estimation has recently been validated in a clinical setting against the TREB of a polysomnography (PSG) setup, with an accuracy of 99.5% (+/- 3 breaths per minute; brpm) and a mean absolute error (MAE) of 0.48 brpm for median RR of 139 whole night recordings.(22) The main objective of the present investigation was to assess the long-term validity of these findings within a home setting by comparing RR estimates with built-in software data in a cohort of patients with domiciliary ventilator support. Within a subset of patients undergoing non-invasive ventilation (NIV) due to COPD, an exploratory analysis was conducted to determine the potential of the CSM in detecting variations in RR preceding impending exacerbations. The identification of such variations could hold implications for clinical applications. Including a second group of patients treated with invasive mechanical ventilation (IMV) aimed to mitigate the negative impact of facemask leakage on the reliability of NIV data.(23) Considering both patient experience and operability measures, the present study is the first to demonstrate the feasibility of this CSM for valid long-term home RR monitoring.

## METHODS

### Patients

Between September 2020 and May 2022, a total of 19 patients receiving NIV therapy (“NIV patients”) were included at the study hospital (University Medicine Essen, Ruhrlandklinik, Germany). Additionally, seven patients treated with IMV were recruited from a collaborating outpatient critical care facility (“IMV patients”). All patients or their legal representatives were fully informed about the study and provided written informed consent upon being given adequate time to consider participation. The study protocol was approved by the ethics committee of the University of Duisburg-Essen (19-8961-BO) and was performed in accordance with the Declaration of Helsinki.

Patients had to be ≥18 years of age and showed the willingness and the ability to comply with the study protocol. Owing to the exploratory nature of the present study, patient selection based on the availability of eligible long-term NIV users at the study hospital, with focus on COPD patients. Gravidity or the presence of a pacemaker were considered as exclusion criteria. Detailed characteristics of enrolled patients are shown in Table 1.

**Table 1.**
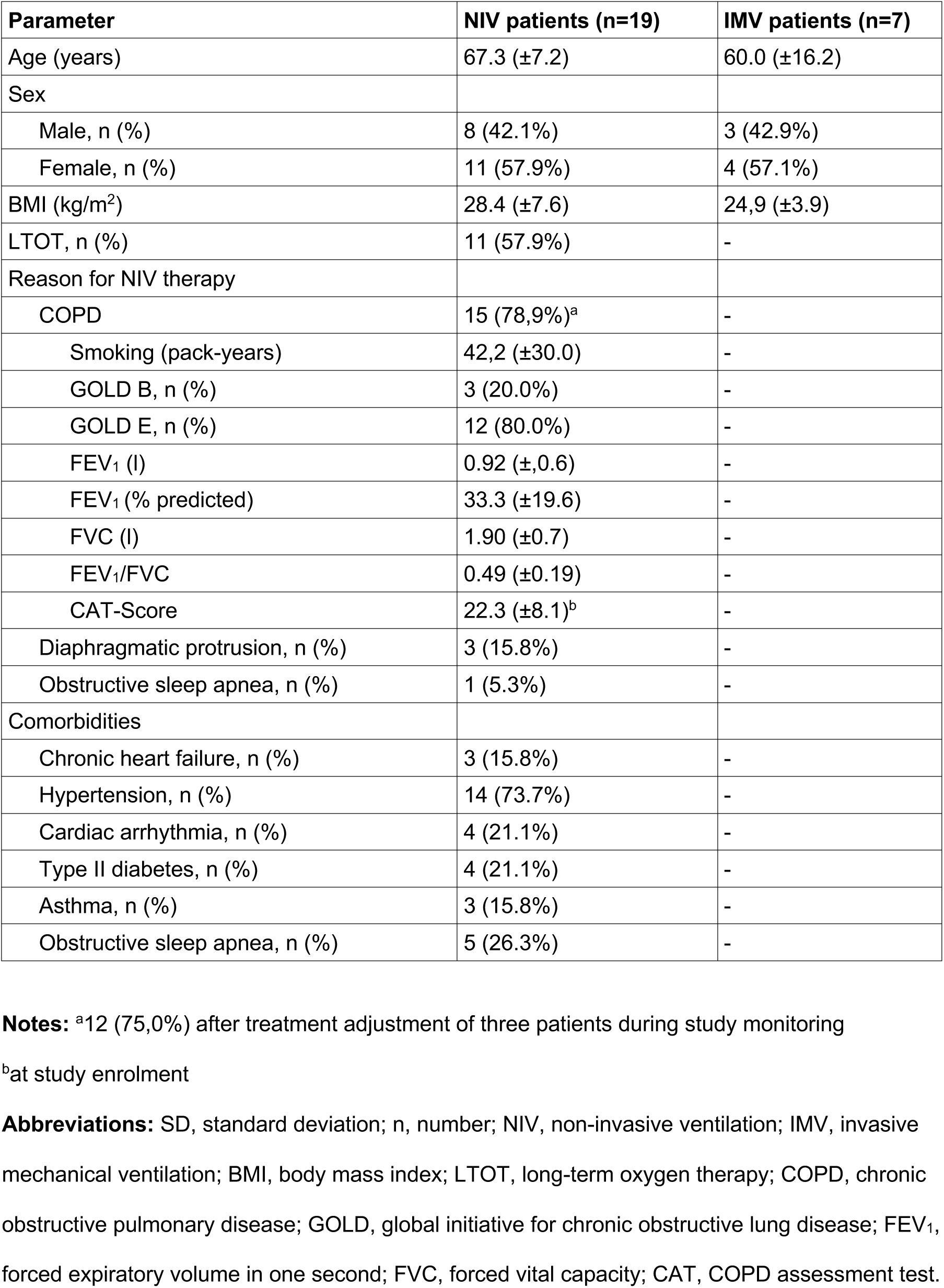
Patient characteristics (data reported as mean ± SD)

| Parameter | NIV patients (n=19) | IMV patients (n=7) |
| --- | --- | --- |
| Age (years) | 67.3 (±7.2) | 60.0 (±16.2) |
| Sex |  |  |
| Male, n (%) | 8 (42.1%) | 3 (42.9%) |
| Female, n (%) | 11 (57.9%) | 4 (57.1%) |
| BMI (kg/m<sup>2</sup>) | 28.4 (±7.6) | 24,9 (±3.9) |
| LTOT, n (%) | 11 (57.9%) | - |
| Reason for NIV therapy |  |  |
| COPD | 15 (78,9%)<sup>a</sup> | - |
| Smoking (pack-years) | 42,2 (±30.0) | - |
| GOLD B, n (%) | 3 (20.0%) | - |
| GOLD E, n (%) | 12 (80.0%) | - |
| FEV<sub>1</sub> (l) | 0.92 (±0.6) | - |
| FEV<sub>1</sub> (% predicted) | 33.3 (±19.6) | - |
| FVC (l) | 1.90 (±0.7) | - |
| FEV<sub>1</sub>/FVC | 0.49 (±0.19) | - |
| CAT-Score | 22.3 (±8.1)<sup>b</sup> | - |
| Diaphragmatic protrusion, n (%) | 3 (15.8%) | - |
| Obstructive sleep apnea, n (%) | 1 (5.3%) | - |
| Comorbidities |  |  |
| Chronic heart failure, n (%) | 3 (15.8%) | - |
| Hypertension, n (%) | 14 (73.7%) | - |
| Cardiac arrhythmia, n (%) | 4 (21.1%) | - |
| Type II diabetes, n (%) | 4 (21.1%) | - |
| Asthma, n (%) | 3 (15.8%) | - |
| Obstructive sleep apnea, n (%) | 5 (26.3%) | - |

**Notes:** <sup>a</sup>12 (75,0%) after treatment adjustment of three patients during study monitoring
<sup>b</sup>at study enrolment
**Abbreviations:** SD, standard deviation; n, number; NIV, non-invasive ventilation; IMV, invasive mechanical ventilation; BMI, body mass index; LTOT, long-term oxygen therapy; COPD, chronic obstructive pulmonary disease; GOLD, global initiative for chronic obstructive lung disease; FEV<sub>1</sub>, forced expiratory volume in one second; FVC, forced vital capacity; CAT, COPD assessment test.

While all 19 patients received NIV therapy at study enrollment, three patients had their therapy modified over time. In two patients, therapy was changed from NIV to long-term oxygen therapy (LTOT) and one patient switched from NIV to continuous positive airway pressure (CPAP) therapy. Conditions for NIV treatment were COPD (n=15), diaphragmatic protrusion (n=3), and severe obstructive sleep apnea (OSA; n=1). Main condition for IMV treatment was respiratory insufficiency due to hypoxic or hemorrhagic brain damage (n=7).

### Study design and experimental procedures

According to the study protocol of this prospective monocentric proof-of-concept trial, nocturnal home RR measurements were expected to be collected for at least six months for NIV patients, and at least one month for IMV patients.

The NIV patients’ hospitalizations, during which they were informed about the experiment, were either scheduled for stationary routine control of NIV-settings or following AECOPD treatment. Upon the patients’ agreement, a PSG was additionally recorded during these nights. The PSG-derived TREB signal was used to assess agreement between built-in software RR estimates of the NIV devices and traditional respiratory effort measures in this patient cohort. Good agreement between these two methods would indirectly support the validity of the CSM RR estimates when compared to NIV data in the home environment.

NIV patients received the CSM after hospital discharge. They were instructed to place it at body height, 40-50 cm from the body next to the bed, directed towards the lower thoracic and upper abdominal region. The transmission area needed to be clear of interfering objects, except for clothing or bedspreads. Patients were advised to turn on the device and associated hotspot only once, as the CSM was programmed for automatic recording between 9 pm and 9 am.

During monthly follow-up calls, COPD patients reported details on doctor visits and episodes of subjective health deterioration for the retrospective identification of potential exacerbation events. AECOPD were defined as periods of significantly higher symptom load that required the use of prescribed rescue medication (oral steroids and/or antibiotics) or led to hospitalization.

Finally, to gather information about the patients’ experiences with the device, a patient-reported experience measurement (PREM) questionnaire was handed out to all 19 NIV patients at the end of the study. The questionnaire comprised 38 items covering different experience categories and has been developed for this device in German language. Categories included handling, acceptance, privacy concerns, benefits and actions they expect from such telehealth programs, and overall satisfaction. Patients also had the opportunity to express problems and suggest improvements through free-text responses.

IMV patients were given the contactless device at the collaborating outpatient critical care facility. As these patients were unable to complete the PREM questionnaire, the caregivers were asked about their experience with the CSM in their daily work routine instead. A flowchart illustrating the study procedure can be found in the online supplemental Figure S1.

### Patient and public involvement

The patients and the public were not involved in the design, conduct, reporting, or dissemination plans of this research. However, the PREM results were intended to serve this very purpose in the future.

### Device specifications

The CSM is able to provide RR estimates in a completely contactless manner. Based on doppler radar technology, it measures thoracic movements with sub-millimeter resolution and uses these to estimate RR via signal processing and machine learning algorithms. Measured data is transmitted daily via hotspot to a cloud server where RR statistics can be monitored by authorized persons, such as the attending physicians or the patients themselves. A more detailed description of technical specifications and algorithmic RR computation can be found in the clinical validation report of this device.(22) To minimize missing data, patients were offered assistance in rebooting the device or the hotspot, as well as optimizing the device positioning as necessary. To be considered for analysis, recordings required a minimum duration of 120 minutes where RR estimates where available. The CSM does not generate RR output when device positioning is poor or when the monitored person is tossing and turning.

Ventilator built-in software data were either collected via a cloud-based telehealth platform or by manual read out using the manufacturers’ software. Ventilator type specifications and quantity are summarized in the online supplemental Table S1.

For PSG data collection, the Nox A1 recording system (Nox Medical, Reykjavík, Iceland) was used.

### Data preprocessing and analysis

In the NIV patient group, the CSM recorded an average of 203 nights per patient with 5.7 hours of suitable contactless data per night, which remained constant throughout the study. NIV data were not available for three patients, resulting in 2326 overlapping nights from 16 patients. Comparison between NIV and TREB data was possible for 12 nights from 10 patients who underwent PSG measurement during the study period, with a maximum of two consecutive nights per patient included.

In the IMV patient group, a total of 334 valid CSM recordings (on average 47 nights per patient) and 505 ventilator datasets were evaluated, resulting in 215 overlapping nights from 4 patients, as three IMV devices failed to store RR data for export. The online supplemental Figure S2 illustrates both the quantity of collected data and the reasons for missing recordings.

Outcome comparison was performed twice, first with time-synchronized data using NIV recordings as a reference, and once without time-synchronization. Synchronization of the time-series data limited the RR analysis of the TREB and the CSM data to the two longest periods of continuous ventilator use overnight. Despite this, the ventilator data represents nightly averages, as detailed assessment for individual time periods was not possible. A minimum valid overlap duration of 60 minutes was set as a requirement for analysis.

Without time-synchronization, RR estimates were analyzed based on device output, irrespective of total recording time or mandatory overlap, reflecting uncontrolled conditions.

RR estimates of interest comprised the 5^th^, 50^th^ (median), and 95^th^ percentile. The comparison of the different measurement techniques based on the mean absolute error (MAE) of these RR statistics. Bland-Altman and scatter plots were used to illustrate their relationship.

Descriptive statistics (frequency, median, mean ± standard deviation) were used for the analysis of the PREM questionnaire. All statistics were performed with IBM SPSS Statistics (Version 28).

## RESULTS

### Respiratory rate statistics

Outcome comparisons between TREB and NIV-software data yielded a MAE of 0.92 ± 1.10 brpm for the 5^th^ percentile of the RR and 0.33 ± 0.47 brpm for the median RR, respectively. However, the analysis revealed rather poor agreement between NIV and TREB for the 95^th^ percentile of the RR with a MAE of 4.25 ± 4.51 brpm (Table 2). The analyses with the time-synchronized data showed similar results for the 5^th^ percentile and for the median, but slightly better agreement for the 95^th^ percentile of the RR with a MAE of 3.57 ± 4.78 brpm (Table 2).

**Table 2.** Respiratory rate statistics comparison.

|  | NIV vs. TREB |  |  | CSM vs. IMV |  |  | CSM vs. NIV |  |  |
| --- | --- | --- | --- | --- | --- | --- | --- | --- | --- |
| RR | 5 % | 50 % | 95 % | 5 % | 50 % | 95 % | 5 % | 50 % | 95 % |
| Without time synchronization |  |  |  |  |  |  |  |  |  |
| n | 10<sup>a</sup> | 12 | 12 | 215 | 215 | 215 | 2116<sup>a</sup> | 2326 | 2326 |
| MAE | 0.92 | **0.33** | 4.25 | 0.44 | **0.12** | 1.90 | 0.43 | **0.78** | 2.85 |
| ± SD | ± 1.10 | ± 0.47 | ± 4.51 | ± 0.63 | ± 0.52 | ± 2.63 | ± 0.84 | ± 1.96 | ± 3.12 |
| With time synchronization |  |  |  |  |  |  |  |  |  |
| n | 10<sup>a</sup> | 12 | 12 | 215 | 215 | 215 | 2008<sup>a</sup> | 2200 | 2200 |
| MAE | 0.84 | **0.38** | 3.57 | 0.44 | **0.12** | 1.90 | 0.36 | **0.65** | 2.81 |
| ± SD | ± 1.17 | ± 0.54 | ± 4.78 | ± 0.63 | ± 0.52 | ± 2.63 | ± 0.65 | ± 1.90 | ± 3.10 |

**Notes:** <sup>a</sup>three NIV devices did not provide 5<sup>th</sup> percentile values,
MAE for 50<sup>th</sup> percentile values in bold numbers.
**Abbreviations:** NIV, non-invasive ventilation; vs., versus; TREB, thoracic respiratory effort belt; CSM, contactless sleep monitor; IMV, invasive mechanical ventilation; RR, respiratory rate; 5%, 5<sup>th</sup> percentile; 50%, 50<sup>th</sup> percentile (=median); 95%, 95<sup>th</sup> percentile; n, number of recordings included; MAE, mean absolute error (breaths per minute); SD, standard deviation.

Results from the IMV group showed excellent agreement for median RR between CSM and IMV data with a MAE of 0.12 ± 0.52 brpm. MAE was 0.44 ± 0.63 brpm and 1.90 ± 2.63 brpm for the 5^th^ percentile and for the 95^th^ percentile, respectively (Table 2). The results did not change with time series synchronization. Bland-Altman analysis and the corresponding scatterplot for IMV and CSM data without time synchronization are displayed in Figure 1(a).

**Figure 1.**
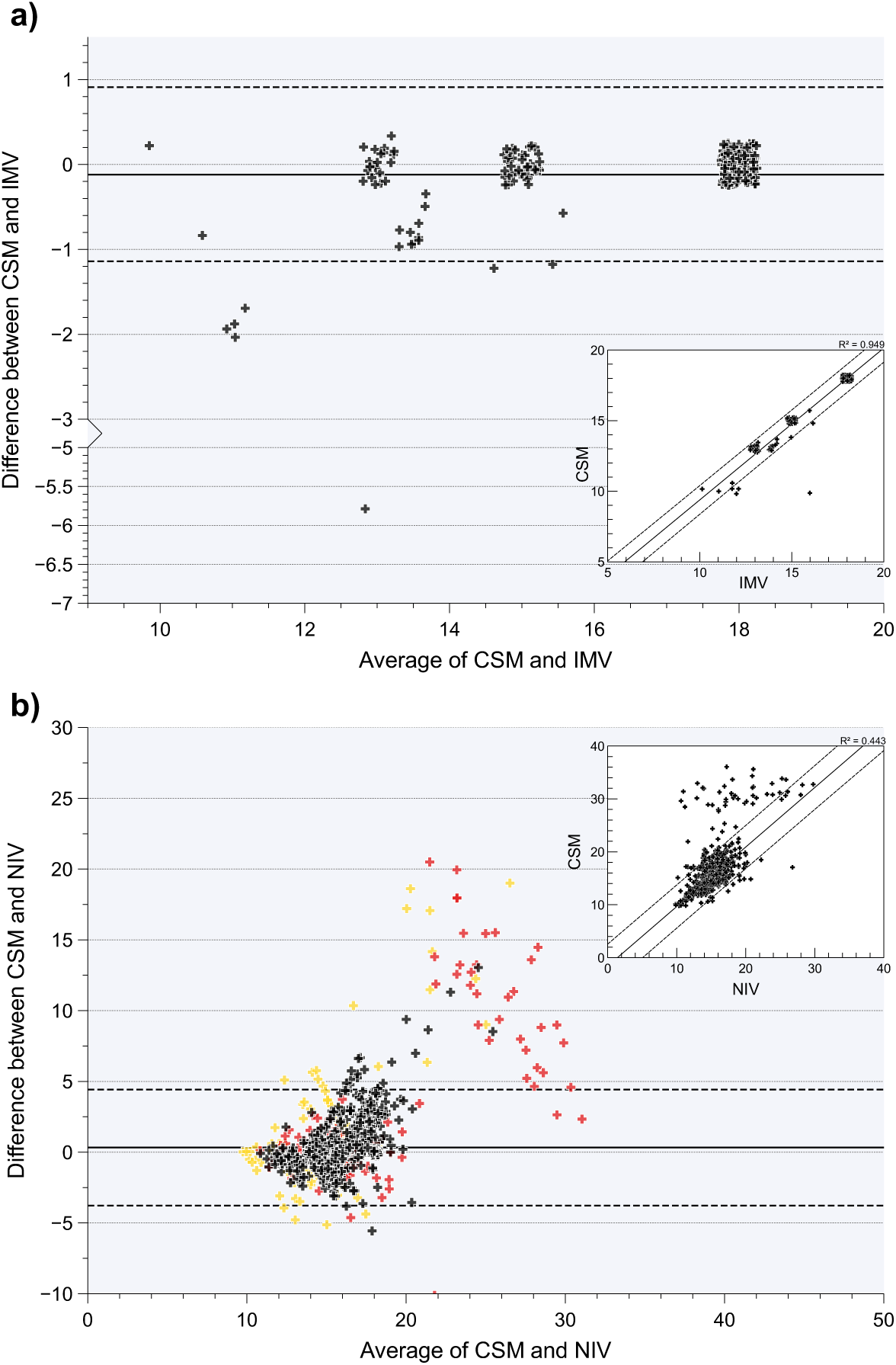
RR comparison between **(a)** CSM and IMV and between **(b)** CSM and NIV [brpm] **Notes:** The main body of the panels **(a, b)** illustrate the Bland-Altman-Analysis. The solid lines indicate the bias (MAE), dashed lines indicate the limits of agreement (mean ± SD*1.96). Additionally, the respective scatter plots are displayed. Jitter has been added in both charts for illustration purposes. The data point clustering in **(a)** is explained by the respiratory backup-rates of the IMV devices. **Abbreviations:** RR, respiratory rate; brpm, breaths per minute; IMV, invasive mechanical ventilator; NIV, non-invasive ventilator; CSM, contactless sleep monitor; MAE, mean absolute error; SD, standard deviation.

Analysis of the home night results derived from the NIV group yielded a MAE for the median RR of 0.78 ± 1.96 brpm. MAE was 0.43 ± 0.84 brpm and 2.85 ± 3.12 brpm for the 5^th^ and for the 95^th^ percentile, respectively (Table 2). When the time-synchronized data based on the NIV usage time were used, the RR statistics comparison showed a marginally higher level of agreement. Bland-Altman analysis and the corresponding scatterplot for data without time synchronization are displayed in Figure 1(b). This figure further highlights that the agreement between the CSM and NIV declines with increasing RR, with higher values detected by the CSM compared to the NIV devices. A closer look on the outliers shows that this is predominantly the case for two patients (colored data points in fFgure 1(b)).

### Patient-reported experience measurement (PREM)

The evaluation of the PREM showed positive results regarding the general usage of the CSM. The overall satisfaction score was 7.9 (± 2.2) out of 10. Mean values for the patient responses within the categories of experience, expectations, concerns, and handling are illustrated in Figure 2. Especially the handling of the device was rated positively, although patients reported that the device sometimes tipped over due to the light construction of the stand base. The potential concerns with the device were strongly dismissed by the patients. IMV patient caregivers indicated that the device was easy to use, although it had to be moved occasionally during nursing procedures. Results for all items are provided in the online supplemental Table S2.

**Figure 2.**
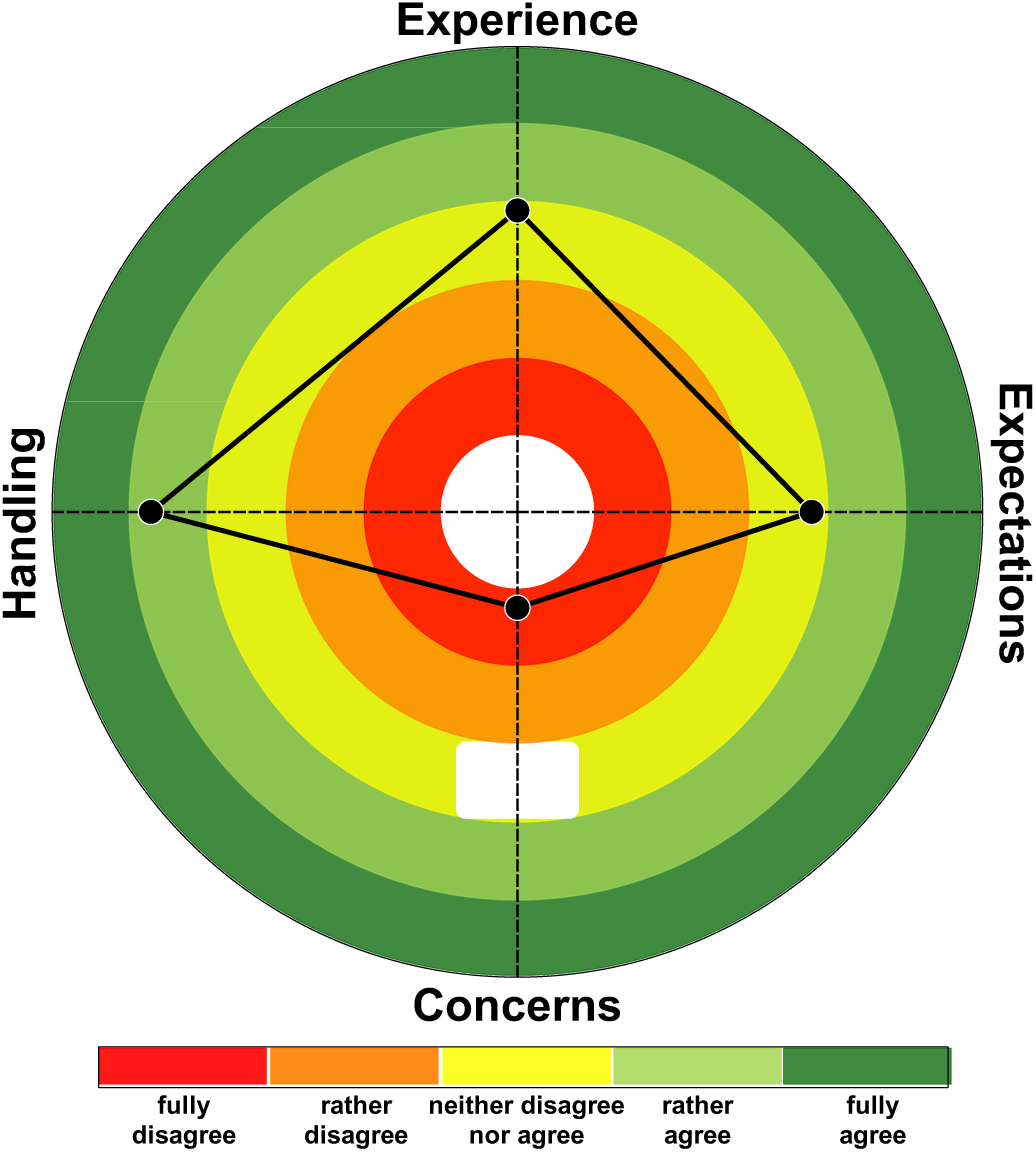
Patient reported experience and operability measures **Notes:** Each point in the polar plot presents the mean PREM response-value for the corresponding category in the NIV group (n=19). In contrast to the other categories, the scale of the category concerns comprised only four points. **Abbreviations:** PREM, patient-reported experience measurement; NIV, non-invasive ventilation.

### Exploring RR in relation to AECOPD

Overall, ten outpatient and three inpatient AECOPD events were observed in a total of six patients. CSM data were unavailable for one AECOPD event, while NIV data were missing for seven cases. Two re-exacerbation events were excluded from analysis as they fell within the guard band of the previous exacerbation. Figure 3 illustrates the median RR measured by the CSM for ten AECOPD events, divided in three time periods: (i) three days before exacerbation onset (prodromal), (ii) three days after exacerbation onset (exacerbation), and (iii) a stable period defined as the nearest consecutive three-day period to the event, which is guarded by a band of seven stable days to another event (baseline). A modest increase in the median RR of 0.75 brpm can be detected during the prodromal phase. As the observed RR variabilities differ notably in terms of their amplitude, onset, and duration between patients, three individual examples of RR time courses related to AECOPD events are given in the online supplemental Figure S3.

**Figure 3.**
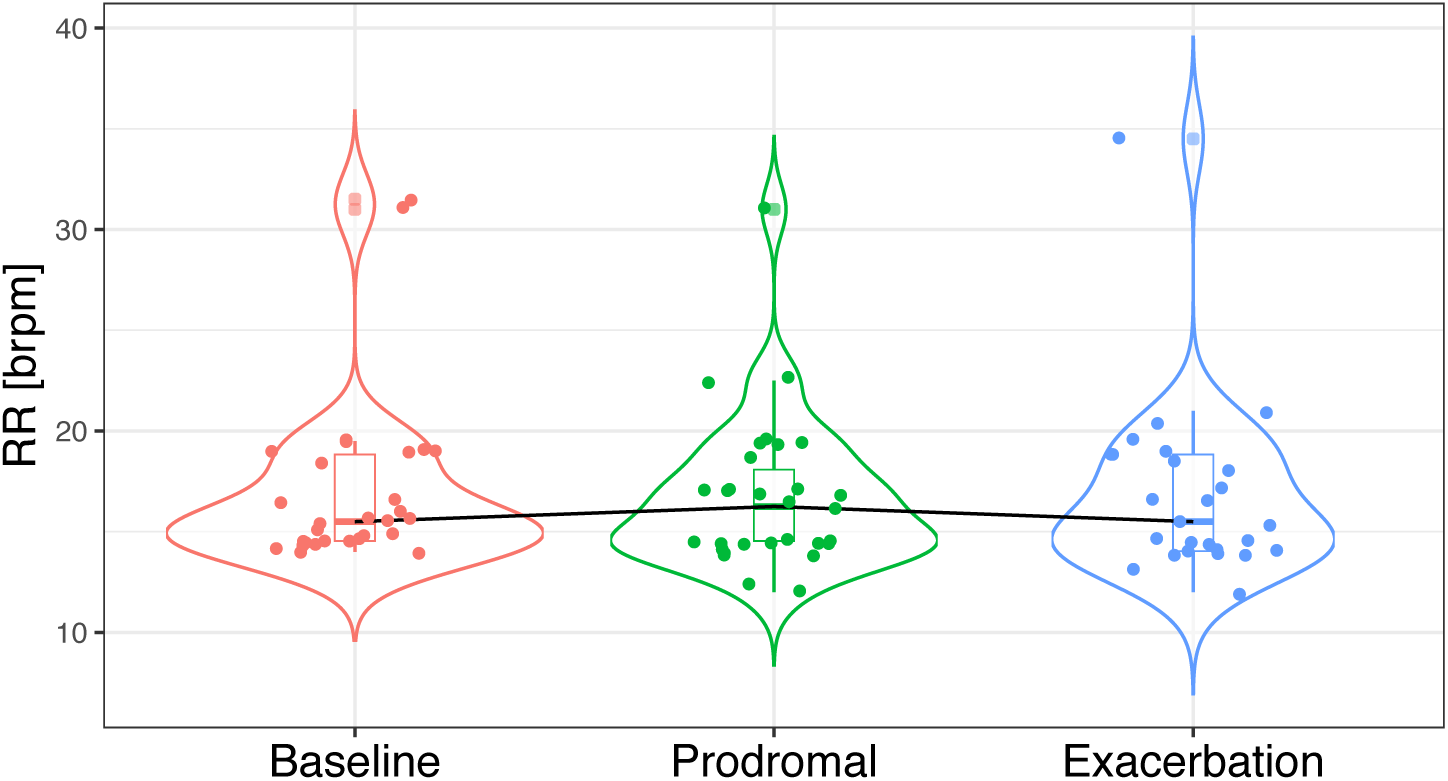
RR assessment in the context of AECOPD events **Notes:** Each violin plot illustrates the distribution of RR across three-day time periods: when symptoms were stable (baseline); three days preceding the event (prodromal); three days starting from the onset day of the exacerbation (exacerbation). Two re-exacerbation events were disregarded as they occurred within the guard band of the previous exacerbation. **Abbreviations:** RR, respiratory rate; brpm, breaths per minute; AECOPD, acute exacerbation of chronic obstructive pulmonary disease.

## DISCUSSION

The present study was the first to measure contactless recorded RR values across multiple nights in the home environment of patients with CRDs requiring additional ventilator support. The findings align with the clinical validation of this device,(22) demonstrating accurate respiratory rate monitoring and favorable patient experiences in the home environment.

As part of the clinical validation, RR estimates were compared with the corresponding outcome of four other contactless respiration monitors. Although the performance of all five sensors were broadly similar, the best agreement was found between the results of the here presented CSM and the SleepMinder/S+ (BiancaMed/ResMed).(22) Unlike the CSM evaluated here, the S+ has had its range of application extended to the home environment without verification of its diagnostic performance in this different setting, and it is not available as medical device.(6)

To the best of our knowledge, only one attempt has been made to validate a novel RR monitor in the home environment. Do *et al* used the TREB signal of a polygraph system as reference to validate a contactless RR monitor at home in a sample of 32 adult and pediatric participants, including healthy volunteers and patients suffering from CRDs.(24) Mean accuracy was 99.0 % (SD: 1.9) and MAE was 0.6 brpm (SD: 0.1) for median RR, which is not much different to the validation results presented here. However, analysis time was restricted to 15 or 120 minutes per patient due to the manual scoring of TREB data. In addition, the study by Do and colleagues covered only one night of monitoring with both measuring methods, rather than multiple nights as in the present investigation. Close to a validation at home, Rubio *et al* compared five minimal-contact home monitoring devices with a vest mounted metabolic sensor system including a facemask in 21 COPD patients during a 57-minute “activities of daily living protocol”.(25) Lowest mean bias and narrowest limits of agreement were found for a chest-band and an accelerometer. In a second step, they tested the acceptability and performance of two selected monitors at home in 23 stable COPD patients during waking hours over 14 consecutive days, but without any further referencing, in contrast to the present study. A common problem with validation studies in the home environment is the selection of an acceptable reference standard that can be used comfortably at home.

For the present study, it was decided to use NIV instead of TREB as a reference to compare long-term CSM RR statistics with an established technology. The rationale for this decision was that (i) integrated NIV software solutions have demonstrated reasonable reliability for most ventilation parameters,(23,26) (ii) previous AECOPD prediction studies have used this measurement method for hypothesis generation,(7,8) and (iii) no other interfering sensors are otherwise required. Moreover, this method is particularly suited for the recording of several consecutive nights, taking both inter- and intraindividual variation into account. However, in addition to these advantages, there are also some issues to consider when applying this approach. Differences in measurements may result from different operation times, since the CSM measures RR continuously regardless of NIV usage. Especially time periods directly before and/or after NIV use may be accompanied by abnormal RR. Therefore, an additional analysis using time synchronization was performed on the data, which was supposed to put this problem into perspective. However, the fact that only average values for NIV and IMV are available for synchronized CSM data poses a limiting factor. Furthermore, data obtained from ventilators can vary across manufactures, and especially the RR and the percentage of cycles triggered by the patient lack validation through bench test studies.(23,26) We addressed this issue by adding a within-subject comparison between NIV and TREB data to the study protocol, offering a specific assessment of the individual reference accuracy for this study collective. The analysis showed sufficient agreement between NIV and TREB for median RR, but revealed poor agreement for the 95^th^ percentile RR, possibly because NIV RR estimates can be compromised by facemask leakage or obstructive events.(23) This possible explanation is firstly supported by the control group results, where the 95^th^ percentile agreement of IMV and CSM was notably higher (Table 2). Secondly, clinical validation showed a MAE for the 90^th^ percentile of 0.67 brpm between CSM and TREB for whole night measurements in the sleep laboratory,(22) which is comparable to the MAE results for the long-term median RR measurements between NIV and CSM. However, the current findings do not support the previous expectation that the performance in the home setting could be superior compared to the in-lab measurement due to favorable measuring conditions Additionally, a comparison between NIV and TREB data was not possible for all patients.

With respect to the acceptability and operability of the CSM, PREM results show that the device can be easily used by a representative group of patients and medical staff. Patients neither felt restricted in their privacy nor bothered by the device. They would appreciate independent access to their data and personal contact to healthcare providers when necessary. Notably, the mobile hotspot used in this study has recently been replaced by a tablet, which provides a communication interface for the patient and thus better research and intervention opportunities. Results for general satisfaction are encouraging, especially when considering the lack of any direct patient advantages in this study. Rubio *et al* reported a few difficulties that patients had with the wearable devices in their study, such as correct positioning on the body or skin problems caused by adhesive patches.(25) Naturally, these issues were not a problem in our study due to the non-intrusive contactless design.

As an exploratory part of our analysis, we assessed RR fluctuations in relation to AECOPD. The averaged RR values show a discrete increase of 0.75 brpm in the median RR prior to an exacerbation. For comparative purposes, the analysis followed Hawthorne *et al*, who observed a 2.0 brpm increase in the median RR during the prodromal phase.(27) In our analysis, we chose to flank the baseline period by a seven-day stable phase similar to the proposal by Shah *et al*.(15) However, individual time courses reveal variations in elevated RR, or no elevations at all, indicating that a personalized assessment should be the aim of future studies to develop robust AECOPD prediction rules. The individual examples of three patients, shown in the online supplemental Figure S3, can be related to already proposed AECOPD prediction rules. The second patient (Figure S3B) shows an elevated RR three days prior to an AECOPD, as well as a higher RR variability in general, which Blouet *et al* considered to be predictive for AECOPD.(7) Interestingly, this severely affected patient accounts for most of the outliers observed in the Bland-Altman analyses (Figure 2, red colored data points), for which it remains speculative which measurement method is more accurate, as the time synchronized analysis showed no differences for these nights. The third patient (Figure S3C) had an increase of 3.0 brpm two days before hospitalization, which is comparable to the findings of Yañez *et al*, who reported that an increase of 2.3 brpm (15 % change from baseline) two days prior to hospitalization was associated with a positive predictive value of 64 % and a negative predictive value of 84 % for AECOPD in their study collective.(9) Although similarities to previous studies can be identified, these parallels should be treated with caution and need to be evaluated by further studies, as the development of a robust prediction rule was not the aim of this proof-of-concept.

Besides RR, oxygen saturation,(28) heart rate,(27) spirometry,(29) and periodic assessment of symptom load are of high interest for the development of AECOPD prediction models.(30) Notably, the CSM evaluated in this study is also able to measure heart rate without contact.

Furthermore, oxygen saturation can now be registered by adding a wireless wearable to preserve the nonintrusive design of this system as best as possible. Symptom questionnaires and patient education programs could be conducted via the tablet. This would provide a unique opportunity to evaluate these indicators in a larger study with one standardized study structure. A long-term goal would be to establish robust parameters and patient-individual prediction rules for AECOPD, including patients at earlier stages of the disease, as remote vital sign monitoring holds the potential to prevent initial exacerbations, a paramount goal in COPD management.(31) A natural progression of this approach would be the evaluation of similar deterioration prediction models in other diseases such as asthma or cystic fibrosis.

## Conclusion

The present study has been the first attempt to examine the long-term validity of contactless recorded RR estimates in the patients’ home environment. The findings are in line with the clinical validation of this device, indicating accurate respiratory rate monitoring accompanied by high patients’ acceptance. Therefore, this study suggests that the device is suitable for future clinical research, focusing on multiparameter approaches and subgroup analyses, as telehealth remains promising in the management of CRD in general and COPD in particular.

## Data Availability

The processed data that support the findings of this study are available from the corresponding author upon reasonable request.

## Abbreviations

AECOPD: acute exacerbation of COPD;
ASB: assisted spontaneous breathing;
BMI: body mass index;
brpm: breaths per minute;
CAT: COPD assessment test;
COPD: chronic obstructive pulmonary disease;
CPAP: continuous positive airway pressure;
CRD: chronic respiratory disease;
CSM: contactless sleep monitor;
FEV_1_: forced expiratory volume in one second;
FVC: forced vital capacity;
GOLD: global initiative for COPD;
IMV: invasive mechanical ventilation;
LTOT: long-term oxygen therapy;
MAE: mean absolute error;
NIV: non-invasive ventilation;
OSA: obstructive sleep apnea;
PREM: patient-reported experience measurement;
PSG: polysomnography;
RR: respiratory rate;
SD: standard deviation;
TREB: thoracic respiratory effort belt;
vs.: versus.

## Ethics statement

This human study was conducted in accordance with the Declaration of Helsinki and was approved by the ethics committee of the University of Duisburg-Essen (19-8961-BO). Each participant or a legal representative gave their written informed consent.

## Acknowledgments

The authors would like to thank all study participants as well as the staff at the sleep laboratory at the study hospital (Ruhrlandklinik, Essen, Germany) and the collaborating outpatient critical care facility (amicu – Außerklinische Intensivpflege, Mühlheim, Germany) for their support in conducting the study. In addition, we appreciate the assistance of Sleepiz AG in the time synchronization and export of RR estimates from the CSM (Python programming language (Python 3.8)).

## Author Contributions

All authors made substantial contributions to the concept of this study, data acquisition and analysis; took part in writing the article or revising it critically; agreed to submit to the current journal and reviewed and agreed to all versions of the manuscript until final publication. All authors agree to be accountable for all aspects of this work.

