## Supplementary material for "At-home validation of remote breathing monitoring: A proof-of-concept for long-term care of respiratory patients using a non-contact, radar-based biomotion sensor": Online Supplemental Figure S1

**SUPLLEMENTAL MATERIAL**

**Figure S1** Flowchart of the study procedures in the NIV- and IMV-group

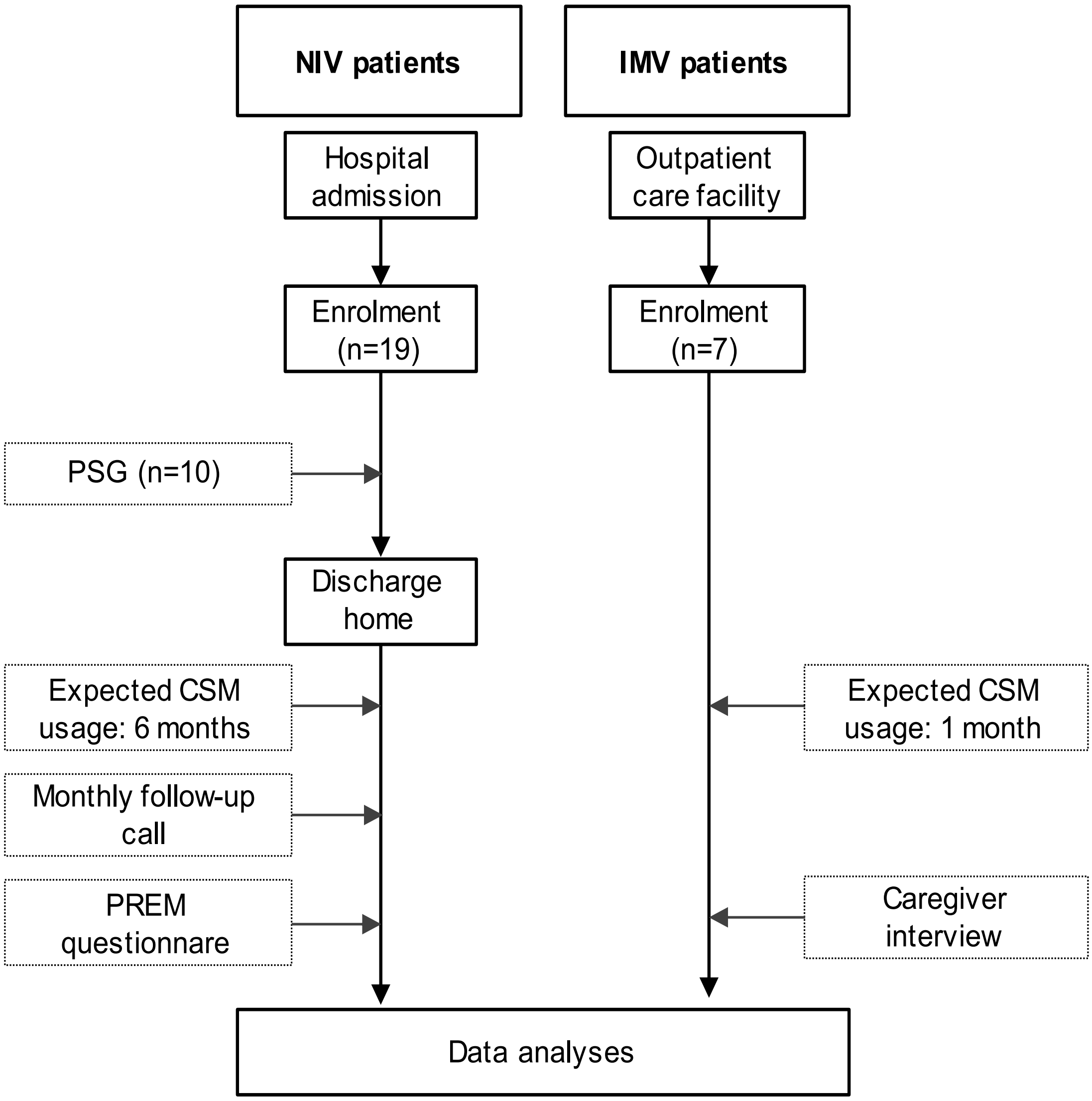


**Abbreviations:** NIV, non-invasive ventilation; IMV, invasive mechanical ventilation; n, number; PSG, polysomnography; CSM, contactless sleep monitor; PREM, patient-reported experience measurement.
