## Supplementary material for "At-home validation of remote breathing monitoring: A proof-of-concept for long-term care of respiratory patients using a non-contact, radar-based biomotion sensor": Online Supplemental Figure S2

**Figure S2** Flowchart of the data collected in the home environment

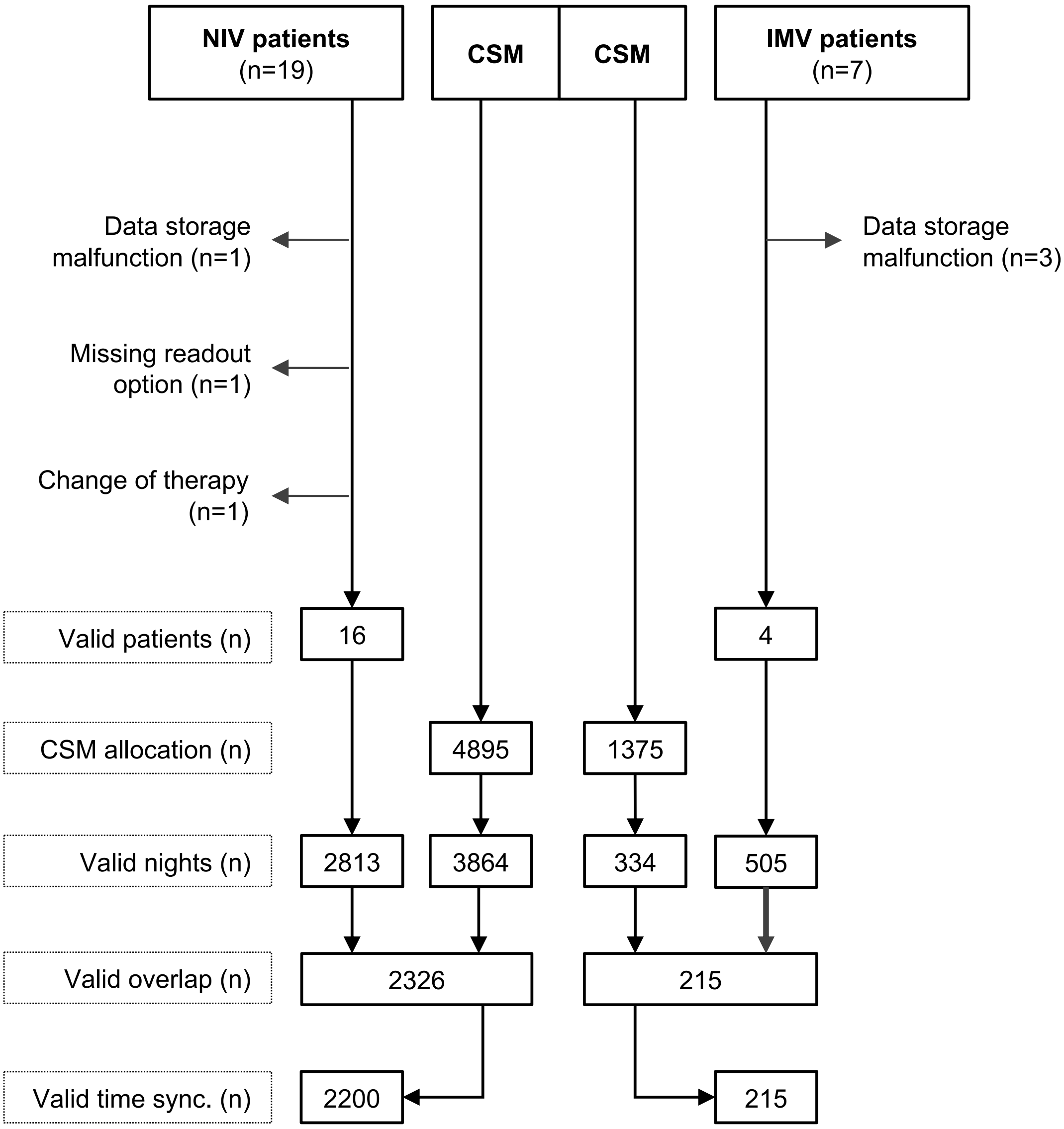


**Notes:** Differences between the allocation duration and valid recordings of the CSM occurred because patients either forgot to switch on the device or the connected hotspot, positioning was suboptimal, or because patients did not use the device, for example, due to hospital admission or delayed start of measurement after recruitment.

**Abbreviations:** NIV, non-invasive ventilation; CSM, contactless sleep monitor; IMV, invasive mechanical ventilation; n, number.
