## Supplementary material for "At-home validation of remote breathing monitoring: A proof-of-concept for long-term care of respiratory patients using a non-contact, radar-based biomotion sensor": Online Supplemental Figure S3

**Figure S3** RR time courses for selected AECOPD events from three different patients (**A, B, C**).

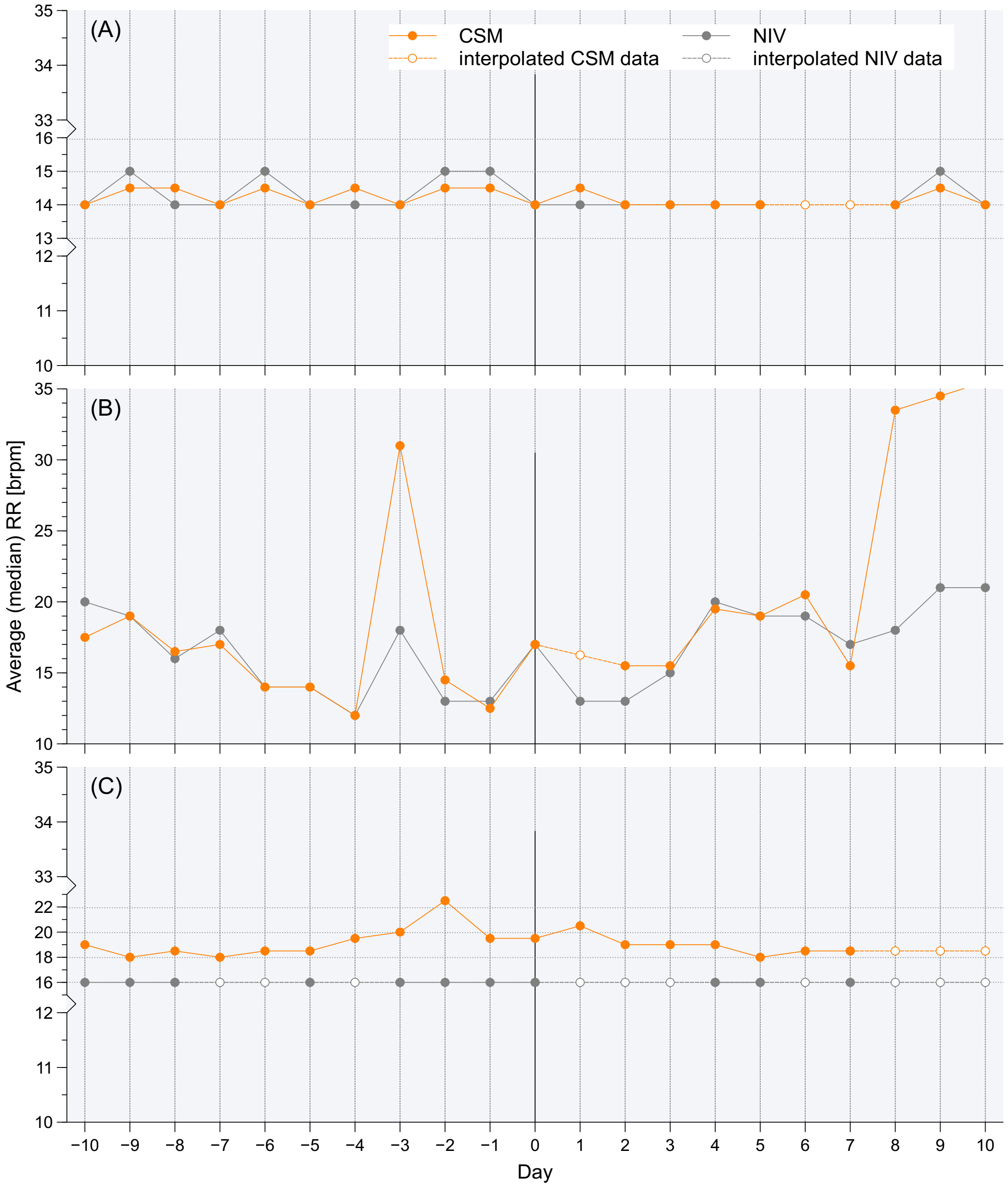


**Notes:** Day zero is defined as the beginning of an AECOPD outpatient treatment or hospital admission. (**A**) shows no significant changes from the baseline regarding an AECOPD event managed on an outpatient basis with corticosteroids. (**B**) displays an increase in the median RR three days prior to the exacerbation event accompanied by overall higher RR variability. (**C**) illustrates an increase two days prior to treatment begin of 3.0 brpm compared to day zero, whereas NIV values indicate no variability at all (NIV in assisted-spontaneous-breathing mode).

**Abbreviations:** RR, respiratory rate; AECOPD, acute exacerbation of chronic obstructive pulmonary disease; NIV, non-invasive ventilator; CSM, contactless sleep monitor; brpm, breaths per minute.
