## Supplementary material for "At-home validation of remote breathing monitoring: A proof-of-concept for long-term care of respiratory patients using a non-contact, radar-based biomotion sensor": Online Supplemental Table S1

**Table S1** Specifications of the ventilators used by the patients.

<table>
<tr><th>n</th><th>Type</th><th>Manufacturer</th><th>Clinical analysis and patient data management software</th><th>Relevant parameters</th></tr>
<tr><td>14</td><td>Stellar 100 / 50</td><td rowspan="3">ResMed Sensor technologies (Dublin, Ireland)</td><td rowspan="3">Cloud-based: AirView,<br><br>PC-based: ResScan</td><td rowspan="2">date, operating hours, RR (median and $5^{th}$ /$95^{th}$ percentile), leakage (median)</td></tr>
<tr><td>7</td><td>Astral</td></tr>
<tr><td>2</td><td>Lumis VPAP</td><td rowspan="2">date, operating hours, RR (median and $95^{th}$ percentile), leakage (median)</td></tr>
<tr><td>1</td><td>Prisma Vent 50</td><td>Löwenstein Medical (Bad Ems, Germany)</td><td>PC-based: PrismaTS</td></tr>
<tr><td>1</td><td>Vigaro Next Generation</td><td>FLO Medizintechnik (Melle, Germany)</td><td>n/a</td><td>n/a</td></tr>
</table>

**Abbreviations:** n, number; RR, respiratory rate; n/a, not available.
