## Supplementary material for "At-home validation of remote breathing monitoring: A proof-of-concept for long-term care of respiratory patients using a non-contact, radar-based biomotion sensor": Online Supplemental Table S2

**Table S2** Means and dispersion of the responses for each item in the PREM questionnaire.

| Category | Mean | SD | P25 | P50 | P75 |
|---|---|---|---|---|---|
| **Experience ("What applies, what does not apply?")**<br>1: fully disagree, 2: rather disagree, 3: neither disagree nor agree, 4: rather agree, 5: fully agree | | | | | |
| I feel / would have felt safer with phone calls | 3,1 | 1,4 | 1,5 | 4,0 | 4,0 |
| I feel safer thanks to the device | 3,0 | 1,5 | 2,0 | 2,0 | 4,0 |
| I feel surveilled and restricted in my privacy | 1,4 | 1,0 | 1,0 | 1,0 | 1,0 |
| Having the device next to my bed bothers me | 1,6 | 1,3 | 1,0 | 1,0 | 1,5 |
| I am satisfied with the device | 4,0 | 0,9 | 4,0 | 4,0 | 5,0 |
| I feel better cared for with my illness | 3,4 | 1,3 | 2,0 | 4,0 | 4,5 |
| I find the design of the device simple | 4,6 | 0,8 | 5,0 | 5,0 | 5,0 |
| My partner is bothered by the device | 2,1 | 0,9 | 1,0 | 2,0 | 3,0 |
| My partner feels disturbed and monitored in their privacy | 2,2 | 1,0 | 1,0 | 3,0 | 3,0 |
| I feel more comfortable undergoing a sleep study at home than in a sleep lab | 4,3 | 1,1 | 4,0 | 5,0 | 5,0 |
| My family is glad that I am being treated to the latest technologies | 3,7 | 1,3 | 3,0 | 4,0 | 5,0 |
| A sleep study at home reduces my anxiety and the stress associated with medical treatment | 3,8 | 1,4 | 3,5 | 4,0 | 5,0 |
| I find the Sleepiz device easier to set up than other devices for sleep testing at home (polygraphy) | 3,7 | 1,1 | 3,0 | 3,0 | 5,0 |
| I sleep better with the Sleepiz device than with other devices for sleep testing at home (polygraphy) | 3,4 | 1,1 | 3,0 | 3,0 | 4,5 |
| **Expectations ("The following conditions should be met for me to use the device permanently")**<br>1: fully disagree, 2: rather disagree, 3: neither disagree nor agree, 4: rather agree, 5: fully agree | | | | | |
| I receive a phone call once a month | 3,8 | 0,7 | 3,3 | 4,0 | 4,0 |
| Someone looks at the data and calls me if necessary | 4,2 | 0,6 | 4,0 | 4,0 | 4,8 |
| I have to see the doctor less often | 3,6 | 1,3 | 4,0 | 4,0 | 4,0 |
| My state of health deteriorates less quickly | 3,9 | 0,8 | 4,0 | 4,0 | 4,0 |
| A deterioration in health / exacerbation can be predicted and prevented | 4,2 | 0,7 | 4,0 | 4,0 | 4,8 |
| I don't have to go to hospital because of a deterioration | 3,7 | 1,1 | 4,0 | 4,0 | 4,0 |
| I can view the collected data myself | 3,6 | 1,0 | 3,3 | 4,0 | 4,0 |
| I can see how I sleep and what other data is recorded by the device | 3,8 | 0,9 | 3,3 | 4,0 | 4,0 |
| My doctor recommends it | 4,1 | 0,5 | 4,0 | 4,0 | 4,0 |

| **Table S2** continued | **Mean** | **SD** | **P25** | **P50** | **P75** |
|---|---|---|---|---|---|
| I can place the device on a bedside table rather than on the stand | 3,1 | 1,4 | 2,0 | 3,0 | 4,0 |
| **If the above conditions apply, I would recommend the device to other patients** (1-10) | 8,1 | 1,4 | 7,0 | 8,0 | 9,0 |
| **I am satisfied with the device** (1-10) | 7,9 | 2,2 | 7,0 | 8,0 | 9,5 |
| **I either store the device or turn in off** 1: daily, 2: almost daily, 3: two to three times a week, 4: rarely, 5: never | 3,5 | 1,7 | 1,5 | 4,0 | 5,0 |
| **Concerns ("I either store the device or turn it off, because...")** 1: fully disagree, 2: rather disagree, 3: rather agree, 4: fully agree | | | | | |
| I feel under surveillance | 1,1 | 0,5 | 1,0 | 1,0 | 1,0 |
| I am afraid of the radiation | 1,1 | 0,2 | 1,0 | 1,0 | 1,0 |
| I want to keep my privacy | 1,4 | 0,8 | 1,0 | 1,0 | 1,5 |
| I am sexually active and feel disturbed by the device | 1,6 | 1,0 | 1,0 | 1,0 | 2,0 |
| I am ashamed of my sleeping habits | 1,1 | 0,5 | 1,0 | 1,0 | 1,0 |
| The device does not look nice | 1,2 | 0,5 | 1,0 | 1,0 | 1,0 |
| **Handling** 1: fully disagree, 2: rather disagree, 3: neither disagree nor agree, 4: rather agree, 5: fully agree | | | | | |
| The components are easy to assemble | 4,7 | 0,6 | 5,0 | 5,0 | 5,0 |
| Height adjustment with the stand is easy | 4,7 | 0,6 | 5,0 | 5,0 | 5,0 |
| Positioning the device is simple | 4,7 | 0,5 | 4,5 | 5,0 | 5,0 |
| Switching on the hotspot is easy | 4,5 | 1,0 | 5,0 | 5,0 | 5,0 |
| Switching on the device is easy | 4,9 | 0,5 | 5,0 | 5,0 | 5,0 |

**Notes:** Free translation from German to English language.

**Abbreviations:** PREM, patient-reported experience measurement; SD, standard deviation; P25, 25$^{th}$ percentile; P50, median; P75, 75$^{th}$ percentile.
